# Clinical academics in the NHS: a cross-section study of research engagement during the monkeypox pandemic

**DOI:** 10.1101/2023.04.22.23288968

**Authors:** Y I Wan, M Smuk, R M Pearse, S Paparini, V J Apea, C M Orkin

## Abstract

**Background:** Recruitment and retention of clinical academics in the UK is under threat. Acute clinical crises can increase opportunities for clinical research. We aimed to examine research involvement amongst clinicians working in sexual health and HIV medicine during the monkeypox (mpox) pandemic and identify factors associated with differential research engagement.

**Methods:** We carried out a cross-sectional study between August and October 2022 using anonymised, self-reported data collected via an online survey disseminated worldwide across multiple specialities. We assessed demographic characteristics, research involvement and outputs, workplace setting, involvement with policy work and public health agencies and media. We examined differences by geographical location comparing the UK, the EU and the US.

**Results:** Of a total 139 respondents from the UK, none identified themselves as clinical researchers, compared to 23/210 (11.0%) from the EU and 5/58 (8.6%) from the US. Overall research engagement was lowest in the UK (15.1% vs. EU 36.7% and US 37.9%). In the UK, research activity was greater amongst consultants (19.5% vs. 18.8% doctors-in-training and 4.9% nurses), those aged 35-50 years (19.7% vs. 15.4% <35 and 8.5% >50 years), males (34.3% vs. 7.1% females and 33.3% non-binary), and those who self-identified as White (15.6% vs.13.3% all other). In research-active individuals, measurable research achievements by journal publications or submissions and obtainment of grant funding were significantly higher in older, male, White, consultants. Less disparity across demographic characteristic groups were seen in both the EU and the US compared to the UK reflecting more diversity amongst research-active clinicians in overall research activity. Markers of research achievement were closer to parity in representation across gender and ethnicity, particularly for the EU.

**Conclusions:** Adherence to and evaluation of existing UK-based recommendations to improve the clinical academic pipeline are needed to increase research engagement and diversity to safeguard UK clinical research in future.

**Key messages:** *What is already known on this topic – summarise the state of scientific knowledge on this subject before you did your study and why this study needed to be done:* The future of clinical academia in the UK is under threat due to a fragile NHS workforce, infrastructure, and environment. Reasons for poor recruitment and retention include lack of mentorship, insufficient job security, delayed career progression, and pay.

*What this study adds – summarise what we now know as a result of this study that we did not know before:* During the mpox pandemic which was an opportunity to produce research, both rates of overall self-reported research engagement and diversity amongst research-active clinicians were significantly lower in the UK compared to both the EU and the US. Reduced engagement with clinical research was especially noticeable in at earlier stages of training, in women, and those from ethnic minority backgrounds.

*How this study might affect research, practice or policy – summarise the implications of this study:* Evaluation of the existing UK-based recommendations to improve the clinical academic pipeline is needed to determine their usefulness. This evaluation should be co-designed by a diverse range of people with protected characteristics with potential to form the future clinical academic pipeline such as junior and senior clinical academics and research-active clinicians.

## Introduction

Clinical academics sit at the interface between research and healthcare, bringing their clinical knowledge into research and exporting their novel therapies, devices and discoveries into clinical care. This synergy can improve clinical outcomes and contribute to efficiencies. In the UK NHS setting, the value of wider engagement with research was clearly evidenced during the COVID-19 pandemic. The UK-led RECOVERY trial was ground-breaking and identified four potential therapies for SARS-CoV-2.^1^ Its delivery was an enormous feat made possible only through a functioning and coordinated network of clinical academic and other research-active NHS staff. It exemplifies the academic power that can be leveraged when cutting-edge academic experts are combined with a large NHS workforce. The availability of an academic clinical pathway may also add interest to medical careers and mitigate the dwindling medical workforce.

However, the number of clinical academics in the UK has declined and there are ongoing disparities in the academic workforce with respect to age, gender, and ethnicity.^2-4^ Unlike the job security of permanent NHS posts, academic posts are more insecure because early career contracts are based on insecure short-term funding with a requirement to continue to get funding.^5^ Additionally, academic training pathways are longer and therefore pay scales increase more slowly. This has contributed to a ‘leaky’ pipeline for young clinical academics who are in their early 30’s. This is particularly applicable to women and others in whom job security is of paramount importance due to financial precarity. These are clear disincentives to pursuing clinical academia as a career path highlighted in a recent inquiry into clinical academic training pathways in the NHS led by Baroness Brown of Cambridge.^6^ Current research is lacking quantitative data on career progression, publication rates and grant successes for clinical academics.

In May 2022, simultaneous human monkeypox (mpox) outbreaks began in Europe and were declared a public health emergency of international concern (PHEIC) by the World Health Organisation (WHO) in June.^7^ The infection almost exclusively affected the networks of gay and bisexual men who have sex with men (GBMSM) and sexual health physicians formed the vanguard of the mpox response in the UK.^8^ The sexual health workforce is comprised of 498 consultants, of which 328 (66%) are female.^9^ Like the COVID-19 pandemic, this represented an enormous challenge to an already stretched sexual health workforce and also a prime opportunity for clinical academics and research-active clinicians to produce high-quality, much-needed clinical research to address the challenge of a re-emerging infection behaving very differently. In this study, we aimed to examine levels of research engagement within a well-defined clinical specialty during a distinct time period. We aimed to determine factors associated with differential rates of research activity in order to assess patterns, trends and potential biases, and better understand reasons and identify solutions for the declining academic workforce.

## Methods

We conducted an international cross-sectional study between August and October 2022 examining engagement with clinical research amongst healthcare professionals involved in the response to the mpox pandemic. To focus on individuals within the sexual health and HIV medicine speciality, we included all individuals who confirmed clinical involvement with the mpox response and had clinical contact with patients in sexual health clinics or HIV clinics. This analysis was restricted to individuals residing in the United Kingdom (UK), the European Union (EU) and the United States (US).

### Data collection

Anonymised, self-reported data was collected via an online survey containing a range of questions on demographic characteristics, involvement in mpox clinical, research, and policy-related work, self-assessment of knowledge and confidence around mpox diagnosis and management and views on outbreak preparedness, educational resources, workload, assessment of risk, and perceptions of moral distress and moral injury. All survey questions examined in this analysis are listed as part of the complete survery in Supplementary materials (S1). The survey was disseminated in English, Spanish, French and Portuguese via the international collaboration Share-Net, an informal network established and led by academic researchers within the London-based Sexual Health and HIV All East Research Collaborative. The survey was disseminated through newsletters and twitter feeds of the British Association for Sexual Health, The British HIV Association, the European AIDS Clinical Society, the International AIDS Society and the research networks of SHARE-net collaborators from 16 countries.

### Ethical approval and regulations

The survey was administered via a survey platform compliant with general data protection regulations (SMART Survey LTD, Tewkesbury, UK) and received ethical approval from the Queen Mary University of London Ethics of Research Committee (QMERC22.297, 27/09/2022). The survey opening page contained information about the aims of the study and custodianship and use of study data. The survey was piloted by ten sexual health clinicians. By clicking ‘continue’ and commencing the survey, individuals were considered to have given consent. Once the survey was closed, partially responded questionnaires were excluded from analysis.

### Statistical analysis

We present descriptive statistics comparing individuals who reported involvement with mpox research and those who did not. We examined demographic characteristics (job title, age, gender, and ethnicity), workplace setting, policy and public health agency work, media engagement, research outputs (publications and grants), role within the research process, and impact on other research responsibilities. Ethnicity was defined using nine categories including a free text category. Due to small numbers within all subgroups apart from White, we report all other groups for this analysis collectively. We examined differences by geographical location comparing the UK, the EU and the US. All analyses were performed using R software v4.02. Results are presented as frequency (percentage): n (%).

## Results

Of a total 139 respondents from the UK, none identified themselves specifically as clinical researchers. Compared to 210 respondents from the EU of whom 23 (11.0%) identified as clinical researchers (19 consultants, 4 doctors in training) and 58 respondents from the US of whom 5 (8.6%) identified as clinical researchers (4 consultants, 1 nurse). Summary characteristics of all included survey respondents are detailed in Supplementary materials (S2: Table S1).

### Research involvement amongst UK clinicians

Amongst UK clinicians, 21 (15.1%) contributed to monkeypox research in any capacity either as an independent researcher, collaborator or contributor. Summary statistics of examined characteristics by job title, age, gender, and ethnicity are detailed in short form in Table 1 and in full in Supplementary materials (S2: Table S2). Of those who contributed to research, the majority (57.1%) reported that due to mpox research, their other research commitments had been affected negatively. More than half (52.4%) published or submitted any research to a scientific journal, over a third (38.1%) were asked to be involved with media outlets, however only one individual (4.8%) obtained grant funding for monkeypox research. Of those that published or submitted research, the majority (63.6%) collected the data and were a named author, but none were involved in study design.

**Table 1.**
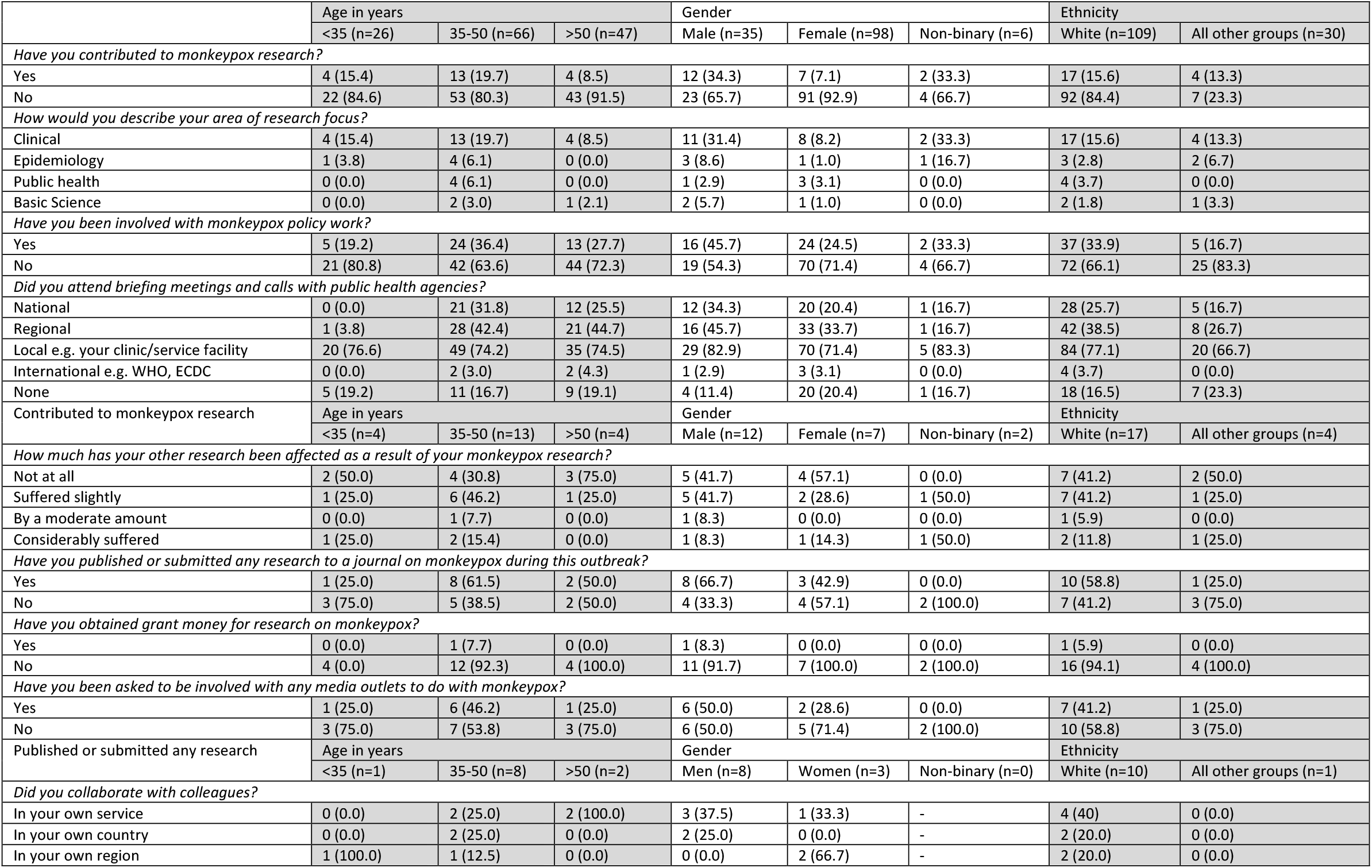
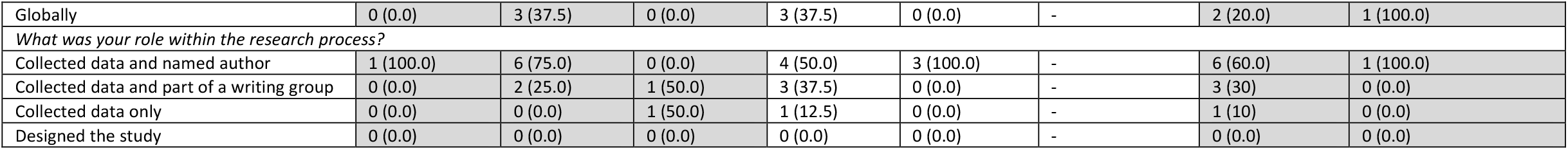
**Short summary of responses to research engagement questions from all UK survey respondents. Total n=139. Categorised by age, gender, and ethnicity. Results presented as n (%). AHP: allied health professional. Full summary table including job title and total columns are detailed in Supplementary materials (S2: Table S2)**.

|  | Age in years |  |  | Gender |  |  | Ethnicity |  |
| --- | --- | --- | --- | --- | --- | --- | --- | --- |
|  | <35 (n=26) | 35-50 (n=66) | >50 (n=47) | Male (n=35) | Female (n=98) | Non-binary (n=6) | White (n=109) | All other groups (n=30) |
| *Have you contributed to monkeypox research?* |  |  |  |  |  |  |  |  |
| Yes | 4 (15.4) | 13 (19.7) | 4 (8.5) | 12 (34.3) | 7 (7.1) | 2 (33.3) | 17 (15.6) | 4 (13.3) |
| No | 22 (84.6) | 53 (80.3) | 43 (91.5) | 23 (65.7) | 91 (92.9) | 4 (66.7) | 92 (84.4) | 7 (23.3) |
| *How would you describe your area of research focus?* |  |  |  |  |  |  |  |  |
| Clinical | 4 (15.4) | 13 (19.7) | 4 (8.5) | 11 (31.4) | 8 (8.2) | 2 (33.3) | 17 (15.6) | 4 (13.3) |
| Epidemiology | 1 (3.8) | 4 (6.1) | 0 (0.0) | 3 (8.6) | 1 (1.0) | 1 (16.7) | 3 (2.8) | 2 (6.7) |
| Public health | 0 (0.0) | 4 (6.1) | 0 (0.0) | 1 (2.9) | 3 (3.1) | 0 (0.0) | 4 (3.7) | 0 (0.0) |
| Basic Science | 0 (0.0) | 2 (3.0) | 1 (2.1) | 2 (5.7) | 1 (1.0) | 0 (0.0) | 2 (1.8) | 1 (3.3) |
| *Have you been involved with monkeypox policy work?* |  |  |  |  |  |  |  |  |
| Yes | 5 (19.2) | 24 (36.4) | 13 (27.7) | 16 (45.7) | 24 (24.5) | 2 (33.3) | 37 (33.9) | 5 (16.7) |
| No | 21 (80.8) | 42 (63.6) | 44 (72.3) | 19 (54.3) | 70 (71.4) | 4 (66.7) | 72 (66.1) | 25 (83.3) |
| *Did you attend briefing meetings and calls with public health agencies?* |  |  |  |  |  |  |  |  |
| National | 0 (0.0) | 21 (31.8) | 12 (25.5) | 12 (34.3) | 20 (20.4) | 1 (16.7) | 28 (25.7) | 5 (16.7) |
| Regional | 1 (3.8) | 28 (42.4) | 21 (44.7) | 16 (45.7) | 33 (33.7) | 1 (16.7) | 42 (38.5) | 8 (26.7) |
| Local e.g. your clinic/service facility | 20 (76.6) | 49 (74.2) | 35 (74.5) | 29 (82.9) | 70 (71.4) | 5 (83.3) | 84 (77.1) | 20 (66.7) |
| International e.g. WHO, ECDC | 0 (0.0) | 2 (3.0) | 2 (4.3) | 1 (2.9) | 3 (3.1) | 0 (0.0) | 4 (3.7) | 0 (0.0) |
| None | 5 (19.2) | 11 (16.7) | 9 (19.1) | 4 (11.4) | 20 (20.4) | 1 (16.7) | 18 (16.5) | 7 (23.3) |
| Contributed to monkeypox research | Age in years |  |  | Gender |  |  | Ethnicity |  |
|  | <35 (n=4) | 35-50 (n=13) | >50 (n=4) | Male (n=12) | Female (n=7) | Non-binary (n=2) | White (n=17) | All other groups (n=4) |
| *How much has your other research been affected as a result of your monkeypox research?* |  |  |  |  |  |  |  |  |
| Not at all | 2 (50.0) | 4 (30.8) | 3 (75.0) | 5 (41.7) | 4 (57.1) | 0 (0.0) | 7 (41.2) | 2 (50.0) |
| Suffered slightly | 1 (25.0) | 6 (46.2) | 1 (25.0) | 5 (41.7) | 2 (28.6) | 1 (50.0) | 7 (41.2) | 1 (25.0) |
| By a moderate amount | 0 (0.0) | 1 (7.7) | 0 (0.0) | 1 (8.3) | 0 (0.0) | 0 (0.0) | 1 (5.9) | 0 (0.0) |
| Considerably suffered | 1 (25.0) | 2 (15.4) | 0 (0.0) | 1 (8.3) | 1 (14.3) | 1 (50.0) | 2 (11.8) | 1 (25.0) |
| *Have you published or submitted any research to a journal on monkeypox during this outbreak?* |  |  |  |  |  |  |  |  |
| Yes | 1 (25.0) | 8 (61.5) | 2 (50.0) | 8 (66.7) | 3 (42.9) | 0 (0.0) | 10 (58.8) | 1 (25.0) |
| No | 3 (75.0) | 5 (38.5) | 2 (50.0) | 4 (33.3) | 4 (57.1) | 2 (100.0) | 7 (41.2) | 3 (75.0) |
| *Have you obtained grant money for research on monkeypox?* |  |  |  |  |  |  |  |  |
| Yes | 0 (0.0) | 1 (7.7) | 0 (0.0) | 1 (8.3) | 0 (0.0) | 0 (0.0) | 1 (5.9) | 0 (0.0) |
| No | 4 (0.0) | 12 (92.3) | 4 (100.0) | 11 (91.7) | 7 (100.0) | 2 (100.0) | 16 (94.1) | 4 (100.0) |
| *Have you been asked to be involved with any media outlets to do with monkeypox?* |  |  |  |  |  |  |  |  |
| Yes | 1 (25.0) | 6 (46.2) | 1 (25.0) | 6 (50.0) | 2 (28.6) | 0 (0.0) | 7 (41.2) | 1 (25.0) |
| No | 3 (75.0) | 7 (53.8) | 3 (75.0) | 6 (50.0) | 5 (71.4) | 2 (100.0) | 10 (58.8) | 3 (75.0) |
| Published or submitted any research | Age in years |  |  | Gender |  |  | Ethnicity |  |
|  | <35 (n=1) | 35-50 (n=8) | >50 (n=2) | Men (n=8) | Women (n=3) | Non-binary (n=0) | White (n=10) | All other groups (n=1) |
| *Did you collaborate with colleagues?* |  |  |  |  |  |  |  |  |
| In your own service | 0 (0.0) | 2 (25.0) | 2 (100.0) | 3 (37.5) | 1 (33.3) | - | 4 (40) | 0 (0.0) |
| In your own country | 0 (0.0) | 2 (25.0) | 0 (0.0) | 2 (25.0) | 0 (0.0) | - | 2 (20.0) | 0 (0.0) |
| In your own region | 1 (100.0) | 1 (12.5) | 0 (0.0) | 0 (0.0) | 2 (66.7) | - | 2 (20.0) | 0 (0.0) |
| Globally | 0 (0.0) | 3 (37.5) | 0 (0.0) | 3 (37.5) | 0 (0.0) | - | 2 (20.0) | 1 (100.0) |
| *What was your role within the research process?* |  |  |  |  |  |  |  |  |
| Collected data and named author | 1 (100.0) | 6 (75.0) | 0 (0.0) | 4 (50.0) | 3 (100.0) | - | 6 (60.0) | 1 (100.0) |
| Collected data and part of a writing group | 0 (0.0) | 2 (25.0) | 1 (50.0) | 3 (37.5) | 0 (0.0) | - | 3 (30) | 0 (0.0) |
| Collected data only | 0 (0.0) | 0 (0.0) | 1 (50.0) | 1 (12.5) | 0 (0.0) | - | 1 (10) | 0 (0.0) |
| Designed the study | 0 (0.0) | 0 (0.0) | 0 (0.0) | 0 (0.0) | 0 (0.0) | - | 0 (0.0) | 0 (0.0) |

Overall, the majority of survey respondents were Consultants, aged 35-50 years, self-identified as Cis-female, and White. A comparison of demographic characteristics by research activity is shown in Figure 1. Research contribution (Have you contributed to monkeypox research?) was lowest amongst nurses (4.9% vs. 19.5% of consultants and 18.8% of doctors-in-training), those aged >50 years (8.5% vs. 19.7% of 35-50 years and 15.4% of <35 years), female respondents (7.1% vs. 34.3% of male and 33.3% of non-binary respondents), and those from ethnically diverse backgrounds (13.3% vs. 15.6% White). In those who were research-active, only Consultants and significantly higher proportions of those aged 35-50 years (61.5% vs. 25% <35 years and 50% >50 years), identifying as male (66.7% vs. 42.9% female and 0% non-binary) and White (58.8% vs. 25% all other ethnic groups) published or submitted their work to a journal. Similarly, the only individual who obtained grant funding identified as a White male Consultant aged 35-50 years. Individuals within these demographic characteristic groups were also more likely to have engaged with media outlets related to their research.

**Figure 1.**
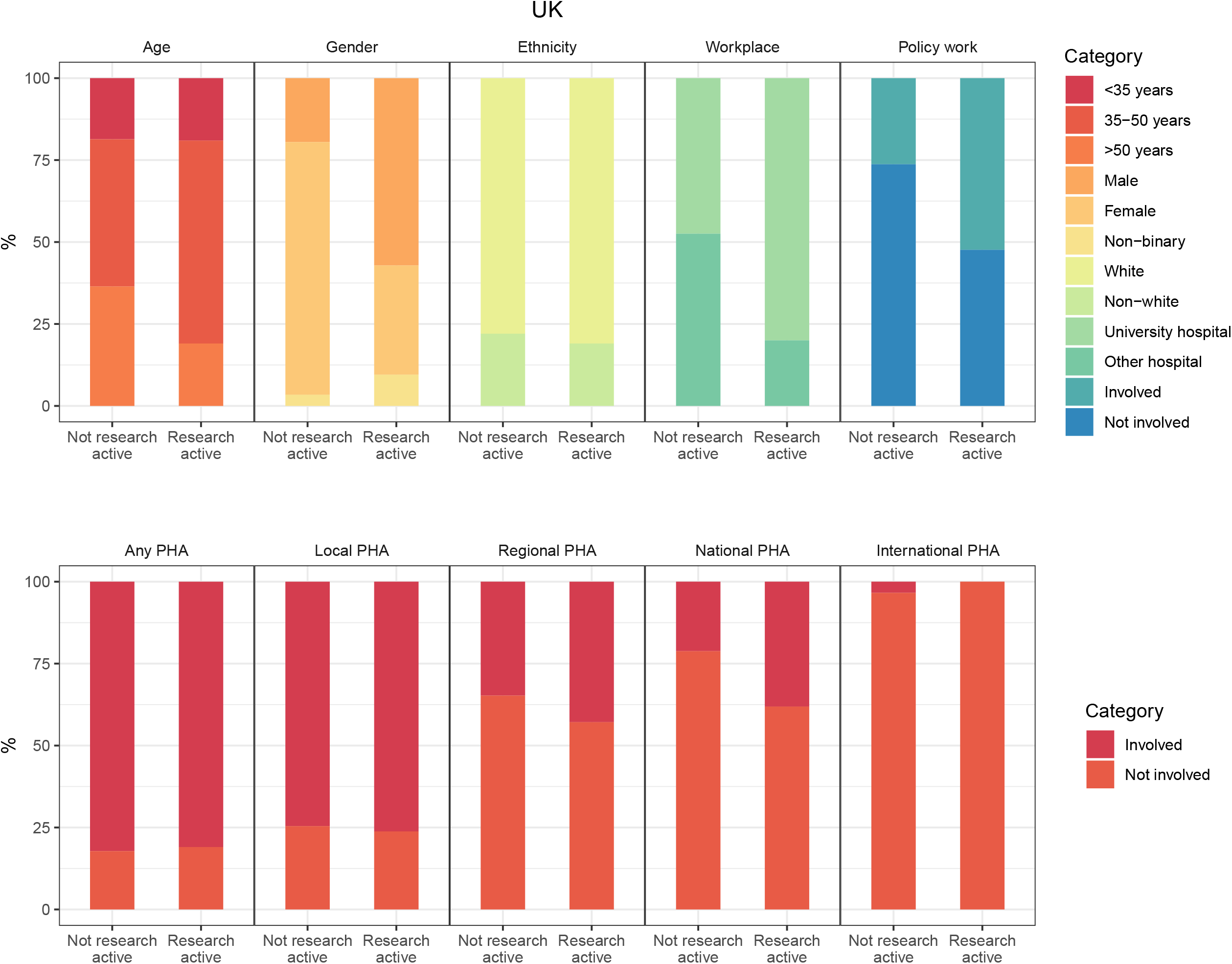
Bar chart comparing research-active and non-research active UK survey participants showing proportions by demographic characteristics, workplace, involvement in policy and public health agency (PHA) work.

Individuals who were research active were more likely to work in a University hospital (80%) compared to those that were not research-active (47.5%). Greater proportions of research-active survey respondents were involved in policy work (52.4% vs. 26.3%). Similar proportions of research active and non-research active respondents had engagement with public health agencies (81.0% vs 82.2%). Most public health agency engagement was at a local level across both groups. By demographics, involvement with both policy work and engagement with public health agencies were highest in those identifying as Consultants, aged 35-50 years, male, and White compared to all other groups.

### Comparison with other geographical regions

Overall research involvement was over two times higher in both the EU (36.7%) and the US (37.9%) compared to the UK. Detailed summary statistics are presented in Supplementary materials (S2: Tables S3 and S4). Compared to the UK, slightly higher proportions of EU clinicians published or submitted their research to a journal (55.8% vs. 52.4%) and were asked to engage with the media (41.6% vs. 38.1%). Two (2.6%) EU respondents obtained grant funding. Although the lowest proportions of US clinicians published or submitted their research to a journal (27.3%), greater proportions engaged with media outlets (45.5%) and obtained grant funding (18.1%). Research active individuals were more likely to be involved in study design in the EU (14.0%) and the US (16.7%) compared to the UK (0%).

Similar to the UK, the majority of survey respondents in both the EU and the US were Consultant-grade equivalents, aged 35-50 years, identified as female, and White. However, amongst those that were research active, less disparity across demographic characteristic groups were seen in both the EU and the US compared to the UK reflecting greater diversity amongst research active healthcare professionals (Figures S1 and S2). Unlike the UK, research activity was higher amongst doctors-in-training (52.9% EU, 50% US) and those aged <35 years (56.4%, 50% US). Higher proportions of ethnically diverse respondents (56.2%) engaged in research than White respondents (35.1%) in the EU. More female (42.3%) respondents than male (33.3%) engaged in research in the US. Similar to the UK, in both the EU and the US, journal publication or submission, involvement with media outlets, and grant funding success were higher amongst Consultants and those aged 35-50 and >50 years. However, there was more equal representation across gender and ethnicity for these markers of research achievements, particularly for the EU.

There were greater differences in percentages of individuals who worked in a University hospital between research active and non-research active individuals in the US (95.5% vs 30.6%) compared to the EU (80.5% vs. 68.4%). In both regions, more research active respondents were also involved in policy work (EU 49.4% vs 29.3%, US 40.9% vs 25%). Similar to the UK, engagement with public health agencies were significant higher in research active compared to non-research active respondents in the EU (81.8% vs. 70.7%) in contrast to the US (18.2% vs. 27.8%). There were greater levels of international public health agency engagement in these regions, particularly in the EU. There was again more equal distribution across demographic characteristic groups in those involved with both policy work and engagement with public health agencies in both the EU and the US compared to the UK.

## Discussion

Our cross-sectional study has examined levels of research engagement within a cohort of clinicians working in sexual health and HIV medicine during the mpox pandemic. Our key findings were that only one in six clinically active healthcare professionals in the UK had any form of research involvement during this acute clinical crisis. This was less than half of those residing in the EU and the US. In this predominantly (70%) female specialty in the UK, levels of research activity were nonetheless significantly higher in older, White and male Consultant grade clinicians compared to all other demographic groups. Measures of research success such as journal publications and obtaining grant funding were also higher for individuals fitting these profiles. Less disparity across demographic characteristic groups were seen in both the EU and the US compared to the UK reflecting increased diversity amongst research active clinicians in overall research activity and markers of research achievements (journal submissions/publications and grant funding).

Our findings support that of other studies conducted over the past five years including a large multicentre led systematic review and primary qualitative analysis of clinical academics in the UK showing poor recruitment and retention rates.^3^ Despite a clear need to continue developing clinical academics, these issues have been overlooked as a priority due to economic pressures and increasing clinical backlogs.^6^ However, continuing this approach is short-sighted as research insights and innovations can support the healthcare system by making it more efficient and help address its current backlog and the other challenges that it faces. This is supported by additional financial benefits such as industry funding and increased recruitment to the medical workforce, and enhanced scientific reputation worldwide for UK scienctists and clinicians.^10^ There are likely to be multifaceted reasons for low research engagement such as the competing demands of academic and clinical workloads, funding pressures and unclear career progression pathways.^3^ Unless this trend is reversed, and new ways of increasing the clinical academic workforce are found, the clinical academic workforce is on course for further decline as there are substantially fewer younger clinical academics to replace those who will retire in the next ten years.^2^

We also found that despite being a female-dominated speciality, individuals working in sexual health and HIV medicine who engaged with research were poorly represented by women, people from minority ethnic backgrounds, and younger age groups. This may imply that the factors that known to negatively influence pursuit of academic medicine at earlier stages of training such as lack of mentorship, insufficient job security, delayed career progression and pay may affect women and people from ethnically diverse backgrounds disproportionately.^11^ Despite an overall increase in women and individuals from minority ethnic backgrounds entering medical school in recent years, disparities continue to exisit with increasing levels of seniority across medicine and within academic medicine specifically.^12-15^. Worryingly, frequently reported reasons for those leaving academic medicine include discrimination and differential opportunities within both the academic and clinical environments.^3^ Only 31% of clinical academics are women and female academics receive only 28% of research funding.^16 17^ 82% of clinical academics identify as White and minority ethnic researchers are less likely to receive research funding.^18^ In addition, maternity status and unequal distribution of labour at home were highlighted as barriers during the COVID-19 pandemic leading to disparities in research activity and publications.^19-21^ Although we observed greater levels of research engagement and diversity in the clinical academic workforce in the EU and US, the US in particular report disparities for women and minority ethnic groups for similar reasons as for the UK.^22-24^

Part of pandemic preparedness is establishing and strengthening academic links with hospitals and early identification of research opportunities. In this study we observed high levels of engagement with policy generation and public health agencies – markers of clinical seniority. Yet despite the overall high percentage of clinicians working in University hospital environments, it appears that opportunities to link with frontline clinicians working at high levels to produce research were missed. Three to five months into the mpox pandemic, few research-active individuals reported research outputs and had obtained grant funding in a rapidly evolving situation where both funding calls and fast-track publications were occurring. This highlights the need to find better ways of supporting clinicians who are and who may wish to be engaged with research. Pertinent to the UK situation, a number of wide-ranging recommendations were made following the recent parliamentary inquiry into clinical academics in the NHS led by Baroness Brown of Cambridge.^6^ Recommendations to funding bodies are to improve career precarity of early career clinical academics by extending contracts. Recommendations for the government are extensive and include mitigations around pay, pension contributions and other conditions. Recommendations to hospitals focus on the importance of academic mentorship. This is particularly challenging in non-University hospital environments and for people from minority ethnic backgrounds for whom few role-models exist. Recommendations to NHS trusts and hospitals are to meet the statutory commitment for consultants to spend 25% of their time on non-clinical work such as research. Ultimately, annual research performance metrics should be devised and reported on annually by integrated care boards to the Department of Health and Social Care. As highlighted by previous studies, these multi-faceted future interventions including those intended to address inequities, require careful evaluation to determine their usefulness.^25-27^ Additionally the involvement of junior academic staff and staff with protected characteristics in co-developing the evaluation of these future interventions is vital.

### Strengths and limitations

This study confirms and adds to the body of evidence to support the declining clinical academic workforce and lack of diversity. It has the added strength of being able to assess a well-defined clinical workforce cohort during a distinct time period to better characterise factors in research engagement whilst reducing potential confounders such as differences in opportunities across different medical specialities. Our study sample captured around one third of the overall sexual health and HIV medicine speciality in the UK and reflect the characteristics of the baseline population. However, within subgroups, there were relatively few participants which limited our ability to assess intersectionality. To our knowledge, this is the first empirical study to assess research activity during the mpox pandemic. We aimed to quantify and objectively assess demographic and environmental factors. However, more qualitative approaches are needed to explore personal experiences in order to better understanding individual barriers and facilitators associated with undertaking research.

### Conclusions

Change is needed to increase the clinical academic pipeline in the UK. Additionally, particular attention needs to be paid to the ongoing disparities in research engagement with respect to age, gender and ethnicity in the UK to safeguard clinical research in the future. Engaging a diverse group of junior clinical academics and research-active clinicians within the NHS not yet on an academic pathway in designing the evaluation of parliamentary recommendations is needed. More research into the barriers and facilitators in people with protected characteristics is needed to better understand the structural barriers to clinical research and to provide more equitable conditions for all clinicians and improve overall recruitment and retention of clinical academics.

## Data Availability

All data produced in the present work are contained in the manuscript.

## Statements and declarations

## Acknowledgements

We would like to thank all the international research collaborators that formed the SHARE-NET group.

## Contributors

YW and CO both conceptualised this work and developed the initial draft. MS, RP, SP and VA provided critical insight, revised, and approved the final manuscript.

### Funding

The authors have not declared any specific grant for this work.

