## Supplemental File 1 for "Clinical academics in the NHS: a cross-section study of research engagement during the monkeypox pandemic"

### HEALTHCARE WORKER EXPERIENCE OF THE MONKEYPOX RESPONSE

**Research Questions:**

1. What have been the experiences and perceptions of international healthcare workers of the Monkeypox response?
2. What has been the impact of the monkeypox response on international healthcare workers?

**Objectives:**

- To assess the clinical experience of healthcare workers during the 2022 multi-country outbreak of monkeypox
- To assess the research experience of healthcare workers during the 2022 multi-country outbreak of monkeypox
- To assess the confidence of healthcare workers' knowledge of monkeypox and identify potential knowledge gaps in clinicians working with monkeypox patients
- To assess the safety of healthcare workers during the 2022 multi-country outbreak of monkeypox
- To assess the preparedness of healthcare workers for the 2022 multi-country outbreak of monkeypox

### EXPERIENCES AND PERCEPTIONS OF HEALTHCARE WORKERS OF THE MONKEYPOX RESPONSE: AN INTERNATIONAL SURVEY

#### SURVEY INFORMATION AND INSTRUCTIONS

You are being invited to complete an anonymous research survey. The survey asks for your opinions and feelings about the current multi-country outbreak of monkeypox, and the impact on you as a healthcare worker. It will take about 15 minutes to complete.

If you do not wish to answer a question, please answer "Prefer not to say". The survey is completely anonymous – we will not ask your personal details and we cannot work out who you are by your answers. You should only take part in this research survey if you are over the age of 18.

The survey is being undertaken by: SHARE collaborative (Queen Mary University of London)

**Key points:**

- Your participation in this survey is entirely voluntary
- You may withdraw at any point during the survey by closing the survey tab
- Please note, once you have filled in the survey you will not be able to withdraw your data as we will not know it was you who completed it

- If you choose to complete this anonymous survey, the information you provide will be analysed by researchers at Queen Mary University of London (QMUL) and used to inform public health responses to new epidemics like monkeypox in the future
- Your answers will be treated confidentially, and the information you provide will not allow you to be identified in any research outputs/publications
- Data from your answers will be held securely in the QMUL data 'Safe Haven' (a secure data repository) for 5 years
- Results may be published in social media, reports and journals, or presented at conferences

If you have any concerns about the manner in which the study was conducted, please contact the researcher(s) responsible for the study, Dr Vanessa Apea, at:.

If you have a complaint which you feel you cannot discuss with the researchers, please contact the QMUL Research Ethics team by, providing details of the study, the QMERC reference number (where possible) and details of your complaint.

If you have concerns about monkeypox symptoms, , you can find further information at https://www.who.int/news-room/fact-sheets/detail/monkeypox.

Queen Mary Ethics of Research Committee reference number: QMERC

If you agree to continue and for us to use the information from this survey, please click on NEXT PAGE button below:

#### SECTION 1: CLINICAL ROLE AND SETTING

1. **Have you been involved with monkeypox clinical work, e.g. diagnosing and treating monkeypox patients and/or their contacts?**
   - o Yes
   - o No
   - o Missing

2. **Have you been involved with monkeypox research in any capacity, e.g. independent researcher, collaborator, contributor?**
   - o Yes
   - o No
   - o Missing

3. **Have you been involved with monkeypox policy work, e.g. guideline writing/ national committees/giving informational talks/writing lay summaries?**
   - o Yes
   - o No

*If no to questions 1 end survey*

4. **What best describes your current role? Please select all that apply.**
   - o Doctor in training
   - o General practitioner/family physician/internist
   - o Sexual Health or HIV physician
   - o Infectious Diseases physician
   - o Coloproctologist/Colo-rectal surgeon
   - o Dermatologist
   - o Paediatrician
   - o Obstetrician/gynaecologist
   - o Nurse or nurse practitioner
   - o Physician's assistant
   - o Counsellor/psychologist
   - o Health promotion worker
   - o Clinical researcher

5. **Do you work at a hospital that is attached to a university?**
   - o Yes
   - o No
   - o Missing

6. **Where did you see suspected or confirmed clinical cases of monkeypox? Please select all that apply.**
   - o Sexual health clinic (community, public, private)
   - o Infectious disease clinic
   - o Emergency department

- HIV clinic
- Dermatology clinic
- General practice
- Rural practice
- In-patient ward
- Obstetrics/Gynaecology clinic or ward
- Paediatric clinic or ward
- Other [please specify]

#### SECTION 2: CLINICAL WORK

7. **During the first four weeks since the first case in your country, on average, what percentage of your work time was focussed on the monkeypox response?**
   - Up to 25%
   - Up to 50%
   - Up to 75%
   - More than 75%

8. **After the first four weeks since the first case in your country, on average, what percentage of your work time was focussed on the monkeypox response?**
   - Up to 25%
   - Up to 50%
   - Up to 75%
   - More than 75%

9. **What tasks have carried out as part of your clinical work? Please tick all that apply**
   - Direct patient care (diagnosis/testing/symptom management/vaccination)
   - Contacting monkeypox patients or their contacts yourself
   - Developing local protocols/operational guidance for your clinic/service
   - Procuring treatment (tecovirimat) for your patients
   - Setting up or working at monkeypox vaccine services
   - Providing data to public health agencies
   - Education
   - Other [please specify]

10. **Has your clinic/service removed other clinical responsibilities to allow you to focus on monkeypox related work**
    - Yes
    - No

11. **During the outbreak of monkeypox did you work?**
    - Longer hours
    - Same hours
    - Shorter hours

12. Which clinical guidelines did your clinic/service follow during the monkeypox outbreak? Please tick all that apply

- o Clinic/local service guidelines
- o National guidelines
- o International guidelines e.g. ECDC, WHO, CDC
- o Guidelines from an infectious disease/sexual health/dermatology society (e.g. IDSA, HIVMA, EACS, BASHH, BHIVA, ASHM, SPILF)
- o Own experience

13. How would you rate your knowledge of how to recognise monkeypox before the outbreak? Please choose the best fit.

- o Had never heard of it
- o Knew where it occurred but not how to recognise it
- o Knew where it occurred and how to recognise it
- o Knew where it occurred how to recognise and how to manage it
- o Have seen and treated a case before prior to this outbreak
- o Have seen many cases prior to this outbreak

14. How confident did you feel managing suspected or confirmed clinical cases of monkeypox at the beginning of the outbreak?

- o Not at all confident
- o A little bit confident
- o Fairly confident
- o Very confident
- o Extremely confident

15. Did you misdiagnose anyone with a monkeypox related rash for other conditions initially?

- o Yes
- o No

*If yes – Go to question 16*
*If no – Go to question 17*

16. If yes - what conditions did you misdiagnose monkeypox as? Please tick all that apply.

- o Chickenpox
- o Disseminated Gonorrhoea
- o Syphilis
- o Herpes
- o Impetigo
- o Drug-induced reaction
- o Hand, Foot and Mouth Disease
- o Molluscum contagiosum

- o Anal fissure
- o Haemorrhoids
- o Anal fistula
- o Other [please specify]

17. How confident do you feel recognising and treating monkeypox now?
    - o Not at all confident
    - o A little bit confident
    - o Fairly confident
    - o Very confident
    - o Extremely confident

18. In your opinion, what quality of care do you think your service has provided to monkeypox patients admitted as in-patient?
    - o Extremely poor
    - o Poor
    - o Average
    - o Good
    - o Excellent
    - o Don't know
    - o No cases so far

19. In your opinion, what quality of care do you think your service has provided to monkeypox patients managed as an out-patient/in the community?
    - o Extremely poor
    - o Poor
    - o Average
    - o Good
    - o Excellent
    - o Don't know
    - o No cases so far

20. Did you have meetings/ward rounds with colleagues in your clinic/service to share information and knowledge about monkeypox?
    - o Yes
    - o No

21. Did you form clinical networks with institutions across your region to share information and knowledge about Monkeypox?
    - o Yes
    - o No

22. In the first four weeks of the outbreak in your country, on average, how many hours a week did you spend on calls or meetings about monkeypox?
    - o 0
    - o 1-2

- 3-5
- 6-10
- >10

23. In the first four weeks of the outbreak in your country, were you expected to attend briefing meetings/calls about monkeypox with any of the following groups? Please tick all that apply.
    - National public health agency
    - Regional public health agency
    - Your clinic/service facility
    - International public health agency, e.g. WHO, ECDC
    - Was not expected to attend table + barchart

24. In the first four weeks of the outbreak in your country, how many hours per week were you personally spending on providing data to public health agencies?
    - <1 hour
    - <2 hours
    - 2-4
    - 4-6
    - >6 hours

#### SECTION 3: SAFETY AT WORK

25. How safe have you felt managing suspected or confirmed clinical cases of Monkeypox?
    - Not at all safe
    - A little bit safe
    - Slightly safe
    - Very safe
    - Extremely safe

26. Did your clinic/service perform a risk assessment of your clinic to ensure staff safety whilst dealing with suspected or confirmed clinical cases of monkeypox?
    - Yes
    - No

27. What kind of PPE (Personal Protective Equipment) does your clinic/service recommend when assessing a patient with monkeypox? Please tick all that apply.
    - Disposable long-sleeved gowns
    - Disposable water-resistant aprons
    - Disposable gloves
    - Disposable shoes or boot covers
    - Respiratory protection – Surgical mask
    - Respiratory protection - Filtering Face Piece Type 3 – with no fit testing completed
    - Respiratory protection - Filtering Face Piece Type 3 – with fit testing completed
    - Respiratory protection – N95 mask – with no fit testing completed
    - Respiratory protection – N95 mask – with fit testing completed

- Eye splash protection (google or visor)
- Other [please specify]

28. Overall, how would you describe PPE availability at your clinic/service?
    - PPE is always available
    - PPE is mostly available
    - PPE is generally available
    - PPE is sometimes available
    - PPE is rarely available

29. Did you receive training on how to appropriately put on or take off PPE?
    - Yes
    - No

30. Did your service's PPE guidance change during the course of the monkeypox response?
    - Yes
    - No

*If yes – Go to question 31*
*If no – Go to question 32*

31. If yes - In your opinion, were these changes communicated in a clear and timely manner?
    - Yes
    - No

32. What other resources were you provided with as part of your clinic/service's Monkeypox response? Please select all that apply.
    - Laboratory diagnostics and sequencing
    - Test kits for lesions
    - Vaccinations
    - Antiviral drugs (e.g. tecovirimat)
    - Disinfectants
    - Deep cleaning of equipment
    - Other [please specify]

33. How do you rate the adequacy of the infection control precautions for monkeypox within your clinical service?
    - Entirely adequate
    - Mostly adequate
    - Somewhat adequate
    - Slightly adequate
    - Not at all adequate

34. How at risk do you feel of contracting monkeypox?
    - Not at all at risk

- Slightly at risk
- Somewhat at risk
- Moderately at risk
- Extremely at risk

35. How concerned are you about the risk to your family members/support network of contracting monkeypox?
   - Not at all concerned
   - Slightly concerned
   - Somewhat concerned
   - Moderately concerned
   - Extremely concerned

36. Did you contract monkeypox?
   - Yes
   - No

37. Did any of your colleagues get monkeypox?
   - Yes
   - No

*If yes – Go to question 38*
*If no – Go to question 39*

38. If yes - how many?
   - 1-5
   - 6-10
   - 11-15
   - 16-20
   - 20+

39. Did any of your family members living in your household get monkeypox?
   - Yes
   - No

#### SECTION 4: MONKEYPOX VACCINATION

40. Have you had a smallpox vaccine before this current multi-country outbreak of monkeypox?
   - Yes
   - No
   - Not sure

41. Have you been offered a smallpox vaccine (either ACAM2000® and JYNNEOS™) as vaccination for monkeypox?

- Yes
- No

*If yes – Go to question 42*
*If no – Go to question 45*

42. If yes, have you accepted the offer and received the vaccine?
    - Yes
    - No

43. Was the process to receive the vaccine straightforward and clear?
    - Yes
    - No

44. Do you feel you received the vaccine in a timely and equitable manner?
    - Yes
    - No
    - Don't know

45. If you have not been offered a vaccine for monkeypox, would you like one?
    - Yes
    - No
    - Don't know

46. Do you think we should be offering vaccination for monkeypox for all healthcare professionals caring for managing suspected or confirmed clinical cases of Monkeypox?
    - Yes
    - No
    - Don't know

47. Do you think we should be offering vaccination for monkeypox for the people at high risk of monkeypox infection prior to exposure, i.e. pre-exposure prophylaxis?
    - Yes
    - No
    - Don't know

48. Is vaccination occurring for all people at high risk prior to exposure occurring in your country?
    - Yes
    - No
    - Don't know

49. In your opinion, do you think access to vaccine for monkeypox adequate in your country?
    - Entirely adequate
    - Mostly adequate

- Somewhat adequate
- Slightly adequate
- Not at all adequate
- Not applicable as there is no access

#### SECTION 5: PREPAREDNESS

**50. How prepared were you (personally) for the monkeypox outbreak?**
- Not at all prepared
- Slightly prepared
- Somewhat prepared
- Moderately prepared
- Extremely prepared

**51. In your opinion, has your institution provided clear, timely and authoritative information about monkeypox?**
- Strongly agree
- Agree
- Neutral/Neither agree nor disagree
- Disagree
- Strongly agree

**52. Have you completed any general outbreak management education and training?**
- Yes
- No

**53. Have you received specific education, training or instruction about monkeypox within your facility?**
- Yes
- No

*If yes – Go to question 54*
*If no – Go to question 56*

**54. Did your hospital arrange education, training or instruction? Please select all that apply.**
- In-house practice education
- Lectures, webinars presentations
- Practical Personal Protective Equipment instruction
- Written guidance
- Other [please specify]

**55. How do you rate the adequacy of this education, training, or instruction?**
- Entirely adequate
- Mostly adequate
- Somewhat adequate
- Slightly adequate

- Not at all adequate

56. How satisfied were you with the support that your clinic/service received from your national public health agency, and why?
   - Not at all at risk
   - Slightly at risk
   - Somewhat at risk
   - Moderately at risk
   - Extremely at risk

   Why? ___________________________________

#### SECTION 6: WELLBEING

57. Have you experienced any of the following symptoms due to your work on monkeypox? (This includes both/either clinical and research work) Please tick all that apply.
   - Fatigue
   - Stress
   - Anxiety
   - Emotional distress
   - Depression
   - Other [please specify]

58. Did you experience any of the following symptoms prior to your work on monkeypox? (This includes both/either clinical and research work) Please tick all that apply
   - Fatigue
   - Stress
   - Anxiety
   - Emotional distress
   - Depression
   - Other [please specify]

59. Are your family members/those living with you concerned that you are interacting with/caring for suspected or confirmed clinical cases of monkeypox?
   - Not at all concerned
   - Slightly concerned
   - Somewhat concerned
   - Moderately concerned
   - Extremely concerned

60. How close do you feel to 'burnout' (a state of physical and emotional exhaustion) due to your work on monkeypox? (This includes both/either clinical and research work)
   - Not at all
   - Slight feelings of burnout
   - Moderate feelings of burnout
   - Considerably burnt out
   - Completely burnt out

61. How close did you feel to 'burnout' (a state of physical and emotional exhaustion) prior to your work on monkeypox? (This includes both/either clinical and research work)
   - Not at all
   - Slight feelings of burnout
   - Moderate feelings of burnout
   - Considerably burnt out
   - Completely burnt out

62. During the past 2 years, have you provided clinical care to COVID patients?
   - Yes
   - No

63. Have you heard of the term 'moral distress' before? / Have you heard of the term 'moral injury' before?
   - Yes
   - No

   *Provide definition:*
   **Moral distress** *is defined as the psychological unease generated where professionals identify an ethically correct action to take but are constrained in their ability to take that action. Even without an understanding of the morally correct action, moral distress can arise from the sense of a moral transgression. More simply, it is the feeling of unease stemming from situations where institutionally required behaviour does not align with moral principles. This can be as a result of a lack of power or agency, or structural limitations, such as insufficient staff, resources, training or time. The individual suffering from moral distress need not be the one who has acted or failed to act; moral distress can be caused by witnessing moral transgressions by others.*

   **Moral injury** *can arise where sustained moral distress leads to impaired function or longer-term psychological harm. Moral injury can produce profound guilt and shame, and in some cases also a sense of betrayal, anger and profound 'moral disorientation'. It has also been linked to severe mental health issues.*

64. Does the term moral distress resonate with your experiences at work managing suspected or confirmed clinical cases of monkeypox?
   - Does not resonate at all

- Slightly resonates
- Somewhat resonates
- Moderately resonates
- Extremely resonates

65. Does the term moral injury resonate with your experiences at work managing suspected or confirmed clinical cases of monkeypox?
    - Does not resonate at all
    - Slightly resonates
    - Somewhat resonates
    - Moderately resonates
    - Extremely resonates

66. During the COVID pandemic, have you experienced moral distress/injury in relation to your ability to provide care?
    - Yes
    - No

    Please describe

67. During the monkeypox response, have you experienced moral distress/injury in relation to your ability to provide care?
    - Yes
    - No

    Please describe

68. Thinking specifically about the 12 months before the COVID-19 pandemic (i.e. the year prior to March 2020), did you have experience of moral distress/injury at work?
    - Yes
    - No

    Please describe

69. Has experiencing the Monkeypox outbreak, in addition to the COVID pandemic, made you more or less likely to remain in health as a profession?
    - No change to intent to remain
    - More likely to remain
    - Less likely to remain
    - No change to intent to leave

#### SECTION 7: MONKEYPOX RESEARCH

70. Have you contributed to monkeypox research
    - Yes

- No

*If yes – go to question 71*
*If no – the rest of the questions don't appear go to*

71. How would you describe your area of research focus? (tick all that apply)
    - Clinical
    - Epidemiology
    - Public health
    - Basic Science

72. How much has your other research been affected as a result of your monkeypox research?
    - Not at all
    - Suffered slightly
    - By a moderate amount
    - Considerably suffered
    - Extremely suffered

73. Have you published or submitted any research to a journal on monkeypox during this outbreak?
    - Yes
    - No

*If yes – go to question 74*
*If no – go to question 76*

74. If yes: Did you collaborate with colleagues?
    - In your own clinic/service
    - In your own region
    - In your own country
    - With global collaborators

75. What was your role within the research process?
    - Designed the study
    - Collected Data only
    - Collected data and was a named author
    - Collected data and was part of a writing group

76. Have you obtained grant money for research on monkeypox?
    - Yes
    - No
    - Have applied but have not heard yet

77. Have you been asked to be involved with any media outlets to do with monkeypox?
    - Yes

- No

*If yes – go to question 78*
*If no – go to question 79*

**78. If yes, have you had training?**
- No training at all
- Very little training
- Some training
- A fair amount of training
- A lot of training

**79. If no, have other colleagues from your service/clinic been asked to engage with the media?**
- Yes
- No

#### SECTION 8: DEMOGRAPHICS

**80. Age:**
- 18-25
- 26-30
- 31-34
- 35-40
- 41-50
- 51-60
- 60+

**81. Gender:**
- Cis-Male
- Cis-Female
- Transmale
- Transfemale
- Non-binary/non-conforming
- Prefer not to say

**82. Sexuality: Do you identify as gay or a bisexual man who has sex with men?**
- Yes
- No
- Prefer not to say

**83. WHO region of residence:** (associated countries will be defined)
- European Region
- Region of the Americas
- South-East Asian Region
- African Region

- Eastern Mediterranean Region
- Western Pacific region
- Prefer not to say

**84. Please select country you are working in**

**85. Ethnicity:**

- White/Caucasian
- Black/African American
- Asian/Asian American
- Latinx or Hispanic
- American Indian or Alaska Native
- Middle Eastern or North African
- Native Hawaiian
- Other Pacific Islander
- Mixed or Multiple Ethnic Group
- Other Ethnic Group [please specify]

**86. Do you hold any of the following degrees? Please tick all that apply.**

- Medical Degree (MD or equivalent)
- MSc
- MPH
- PhD
- Other [please specify]

#### SECTION 9: ANY ADDITIONAL COMMENTS?

**87. Any further comments?**

*'Text box should have a maximum of 250 words'*

#### 'THANK YOU' PAGE

Thank you for taking time to take part in our survey and sharing your views. We are grateful for your participation.

IF YOU HAVE ANY FURTHER QUESTIONS ABOUT THIS SURVEY OR IF YOU WOULD LIKE TO TAKE PART IN FUTURE RESEARCH WITH OUR TEAM, PLEASE CONTACT US AT:.
