## Supplemental File 2 for "Clinical academics in the NHS: a cross-section study of research engagement during the monkeypox pandemic"

**Table S1. Characteristics of all included survey respondents. Results presented as n (%). AHP: allied health professional.**

| | UK (n=139) | EU (n=210) | US (n=58) | Total (n=407) |
|---|---|---|---|---|
| *Job title* | | | | |
| Consultant | 82 (59.0) | 178 (84.8) | 40 (69.0) | 300 (73.7) |
| Doctor-in-training | 16 (11.5) | 17 (8.1) | 2 (3.4) | 35 (8.6) |
| Nurse or AHP | 41 (29.5) | 15 (7.1) | 16 (27.6) | 72 (17.7) |
| *Age in years* | | | | |
| <35 | 26 (18.7) | 39 (18.6) | 8 (13.8) | 73 (17.9) |
| 35-50 | 66 (47.5) | 105 (50.0) | 23 ()39.7 | 194 (47.7) |
| >50 | 47 (33.8) | 66 (31.4) | 27 (46.6) | 140 (34.4) |
| *Gender* | | | | |
| Male | 35 (25.2) | 100 (47.6) | 27 (46.6) | 162 (39.8) |
| Female | 98 (70.5) | 106 (50.5) | 30 (51.7) | 234 (57.5) |
| Non-binary | 6 (4.3) | 4 (1.9) | 1 (1.7) | 22 (2.7) |
| *Ethnicity* | | | | |
| White | 109 (78.4) | 194 (92.3) | 38 (65.5) | 341 (83.8) |
| All other groups | 30 (21.6) | 16 (7.6) | 20 (34.5) | 66 (16.2) |

**Table S2. Summary of responses to research engagement questions from all UK survey respondents. Results presented as n (%). AHP: allied health professional.**

| | Job title | | | Age in years | | | Gender | | | Ethnicity | | Total (n=139) |
|---|---|---|---|---|---|---|---|---|---|---|---|---|
| | Consultant (n=82) | Doctor-in-training (n=16) | Nurse or AHP (n=41) | <35 (n=26) | 35-50 (n=66) | >50 (n=47) | Male (n=35) | Female (n=98) | Non-binary (n=6) | White (n=109) | All other groups (n=30) | |
| *Have you contributed to monkeypox research?* | | | | | | | | | | | | |
| Yes | 16 (19.5) | 3 (18.8) | 2 (4.9) | 4 (15.4) | 13 (19.7) | 4 (8.5) | 12 (34.3) | 7 (7.1) | 2 (33.3) | 17 (15.6) | 4 (13.3) | 21 (15.1) |
| No | 66 (80.5) | 13 (81.2) | 39 (95.1) | 22 (84.6) | 53 (80.3) | 43 (91.5) | 23 (65.7) | 91 (92.9) | 4 (66.7) | 92 (84.4) | 7 (23.3) | 118 (84.9) |
| *How would you describe your area of research focus?* | | | | | | | | | | | | |
| Clinical | 18 (22.0) | 3 (18.8) | 0 (0.0) | 4 (15.4) | 13 (19.7) | 4 (8.5) | 11 (31.4) | 8 (8.2) | 2 (33.3) | 17 (15.6) | 4 (13.3) | 21 (15.1) |
| Epidemiology | 4 (4.9) | 1 (6.2) | 0 (0.0) | 1 (3.8) | 4 (6.1) | 0 (0.0) | 3 (8.6) | 1 (1.0) | 1 (16.7) | 3 (2.8) | 2 (6.7) | 5 (3.6) |
| Public health | 2 (2.4) | 0 (0.0) | 2 (4.9) | 0 (0.0) | 4 (6.1) | 0 (0.0) | 1 (2.9) | 3 (3.1) | 0 (0.0) | 4 (3.7) | 0 (0.0) | 4 (2.9) |
| Basic Science | 1 (1.2) | 0 (0.0) | 2 (4.9) | 0 (0.0) | 2 (3.0) | 1 (2.1) | 2 (5.7) | 1 (1.0) | 0 (0.0) | 2 (1.8) | 1 (3.3) | 3 (2.2) |
| *Have you been involved with monkeypox policy work?* | | | | | | | | | | | | |
| Yes | 29 (35.4) | 4 (25.0) | 9 (22) | 5 (19.2) | 24 (36.4) | 13 (27.7) | 16 (45.7) | 24 (24.5) | 2 (33.3) | 37 (33.9) | 5 (16.7) | 42 (30.2) |
| No | 53 (64.6) | 12 (75.0) | 32 (78) | 21 (80.8) | 42 (63.6) | 44 (72.3) | 19 (54.3) | 70 (71.4) | 4 (66.7) | 72 (66.1) | 25 (83.3) | 97 (69.8) |
| *Did you attend briefing meetings and calls with public health agencies?* | | | | | | | | | | | | |
| National | 26 (31.7) | 1 (6.2) | 6 (14.6) | 0 (0.0) | 21 (31.8) | 12 (25.5) | 12 (34.3) | 20 (20.4) | 1 (16.7) | 28 (25.7) | 5 (16.7) | 22 (23.7) |
| Regional | 43 (52.4) | 0 (0.0) | 7 (17.1) | 1 (3.8) | 28 (42.4) | 21 (44.7) | 16 (45.7) | 33 (33.7) | 1 (16.7) | 42 (38.5) | 8 (26.7) | 50 (36.0) |
| Local e.g. your clinic/service facility | 65 (79.3) | 9 (56.2) | 30 (73.2) | 20 (76.6) | 49 (74.2) | 35 (74.5) | 29 (82.9) | 70 (71.4) | 5 (83.3) | 84 (77.1) | 20 (66.7) | 104 (74.8) |
| International e.g. WHO, ECDC | 3 (3.7) | 0 (0.0) | 1 (2.4) | 0 (0.0) | 2 (3.0) | 2 (4.3) | 1 (2.9) | 3 (3.1) | 0 (0.0) | 4 (3.7) | 0 (0.0) | 4 (2.9) |
| None | 9 (11.0) | 7 (43.8) | 9 (22.0) | 5 (19.2) | 11 (16.7) | 9 (19.1) | 4 (11.4) | 20 (20.4) | 1 (16.7) | 18 (16.5) | 7 (23.3) | 25 (18.0) |
| Contributed to monkeypox research | Job title | | | Age in years | | | Gender | | | Ethnicity | | Total (n=21) |
| | Consultant (n=16) | Doctor-in-training (n=3) | Nurse or AHP (n=2) | <35 (n=4) | 35-50 (n=13) | >50 (n=4) | Male (n=12) | Female (n=7) | Non-binary (n=2) | White (n=17) | All other groups (n=4) | |
| *How much has your other research been affected as a result of your monkeypox research?* | | | | | | | | | | | | |
| Not at all | 7 (43.8) | 1 (33.3) | 1 (50.0) | 2 (50.0) | 4 (30.8) | 3 (75.0) | 5 (41.7) | 4 (57.1) | 0 (0.0) | 7 (41.2) | 2 (50.0) | 9 (42.9) |
| Suffered slightly | 6 (37.5) | 1 (33.3) | 1 (50.0) | 1 (25.0) | 6 (46.2) | 1 (25.0) | 5 (41.7) | 2 (28.6) | 1 (50.0) | 7 (41.2) | 1 (25.0) | 8 (38.1) |
| By a moderate amount | 1 (6.3) | 0 (0.0) | 0 (0.0) | 0 (0.0) | 1 (7.7) | 0 (0.0) | 1 (8.3) | 0 (0.0) | 0 (0.0) | 1 (5.9) | 0 (0.0) | 1 (4.8) |
| Considerably suffered | 2 (12.5) | 1 (33.3) | 0 (0.0) | 1 (25.0) | 2 (15.4) | 0 (0.0) | 1 (8.3) | 1 (14.3) | 1 (50.0) | 2 (11.8) | 1 (25.0) | 3 (14.3) |
| *Have you published or submitted any research to a journal on monkeypox during this outbreak?* | | | | | | | | | | | | |
| Yes | 10 (62.5) | 0 (0.0) | 1 (50.0) | 1 (25.0) | 8 (61.5) | 2 (50.0) | 8 (66.7) | 3 (42.9) | 0 (0.0) | 10 (58.8) | 1 (25.0) | 11 (52.4) |
| No | 6 (37.5) | 3 (100.0) | 1 (50.0) | 3 (75.0) | 5 (38.5) | 2 (50.0) | 4 (33.3) | 4 (57.1) | 2 (100.0) | 7 (41.2) | 3 (75.0) | 10 (47.6) |
| *Have you obtained grant money for research on monkeypox?* | | | | | | | | | | | | |
| Yes | 1 (6.3) | 0 (0.0) | 0 (0.0) | 0 (0.0) | 1 (7.7) | 0 (0.0) | 1 (8.3) | 0 (0.0) | 0 (0.0) | 1 (5.9) | 0 (0.0) | 1 (4.8) |
| No | 15 (93.8) | 3 (100.0) | 2 (100.0) | 4 (0.0) | 12 (92.3) | 4 (100.0) | 11 (91.7) | 7 (100.0) | 2 (100.0) | 16 (94.1) | 4 (100.0) | 20 (95.2) |
| *Have you been asked to be involved with any media outlets to do with monkeypox?* | | | | | | | | | | | | |
| Yes | 8 (50.0) | 0 (0.0) | 0 (0.0) | 1 (25.0) | 6 (46.2) | 1 (25.0) | 6 (50.0) | 2 (28.6) | 0 (0.0) | 7 (41.2) | 1 (25.0) | 8 (38.1) |
| No | 8 (50.0) | 3 (100.0) | 2 (100.0) | 3 (75.0) | 7 (53.8) | 3 (75.0) | 6 (50.0) | 5 (71.4) | 2 (100.0) | 10 (58.8) | 3 (75.0) | 13 (61.9) |
| Published or submitted any research | Job title | | | | | | Gender | | | Ethnicity | | Total (n=11) |
| | Consultant (n=10) | Doctor-in-training (n=0) | Nurse or AHP (n=1) | <35 (n=1) | 35-50 (n=8) | >50 (n=2) | Men (n=8) | Women (n=3) | Non-binary (n=0) | White (n=10) | All other groups (n=1) | |
| *Did you collaborate with colleagues?* | | | | | | | | | | | | |
| In your own service | 4 (40) | - | 0 (0.0) | 0 (0.0) | 2 (25.0) | 2 (100.0) | 3 (37.5) | 1 (33.3) | - | 4 (40) | 0 (0.0) | 4 (36.4) |

| | | | | | | | | | | | | |
|---|---|---|---|---|---|---|---|---|---|---|---|---|
| In your own country | 2 (20.0) | - | 0 (0.0) | 0 (0.0) | 2 (25.0) | 0 (0.0) | 2 (25.0) | 0 (0.0) | - | 2 (20.0) | 0 (0.0) | 2 (18.2) |
| In your own region | 1 (10) | - | 1 (100.0) | 1 (100.0) | 1 (12.5) | 0 (0.0) | 0 (0.0) | 2 (66.7) | - | 2 (20.0) | 0 (0.0) | 2 (18.2) |
| Globally | 3 (30) | - | 0 (0.0) | 0 (0.0) | 3 (37.5) | 0 (0.0) | 3 (37.5) | 0 (0.0) | - | 2 (20.0) | 1 (100.0) | 3 (27.3) |
| *What was your role within the research process?* | | | | | | | | | | | | |
| Collected data and named author | 6 (60.0) | - | 1 (100.0) | 1 (100.0) | 6 (75.0) | 0 (0.0) | 4 (50.0) | 3 (100.0) | - | 6 (60.0) | 1 (100.0) | 7 (63.6) |
| Collected data and part of a writing group | 3 (30) | - | 0 (0.0) | 0 (0.0) | 2 (25.0) | 1 (50.0) | 3 (37.5) | 0 (0.0) | - | 3 (30) | 0 (0.0) | 3 (27.3) |
| Collected data only | 1 (10) | - | 0 (0.0) | 0 (0.0) | 0 (0.0) | 1 (50.0) | 1 (12.5) | 0 (0.0) | - | 1 (10) | 0 (0.0) | 1 (9.1) |
| Designed the study | 0 (0.0) | - | 0 (0.0) | 0 (0.0) | 0 (0.0) | 0 (0.0) | 0 (0.0) | 0 (0.0) | - | 0 (0.0) | 0 (0.0) | 0 (0.0) |

**Table S3. Summary of responses to research engagement questions from all EU survey respondents. Results presented as n (%). AHP: allied health professional.**

| | Job title | | | Age in years | | | Gender | | | Ethnicity | | Total |
|---|---|---|---|---|---|---|---|---|---|---|---|---|
| | Consultant (n=178) | Doctor-in-training (n=17) | Nurse or AHP (n=15) | <35 (n=39) | 35-50 (n=105) | >50 (n=66) | Male (n=100) | Female (n=106) | Non-binary (n=4) | White (n=194) | All other groups (n=16) | (n=210) |
| *Have you contributed to monkeypox research?* | | | | | | | | | | | | |
| Yes | 61 (34.3) | 9 (52.9) | 7 (46.7) | 22 (56.4) | 29 (27.6) | 26 (39.4) | 45 (45) | 31 (29.2) | 1 (25.0) | 68 (35.1) | 9 (56.2) | 77 (36.7) |
| No | 117 (65.7) | 8 (47.1) | 8 (53.3) | 17 (43.6) | 76 (72.4) | 40 (60.6) | 55 (55) | 75 (70.8) | 3 (75.0) | 126 (64.9) | 7 (43.8) | 133 (63.3) |
| *How would you describe your area of research focus?* | | | | | | | | | | | | |
| Clinical | 56 (31.5) | 9 (52.9) | 5 (33.3) | 21 (53.8) | 25 (23.8) | 24 (36.4) | 40 (40) | 29 (27.4) | 1 (25.0) | 61 (31.4) | 9 (56.2) | 70 (33.3) |
| Epidemiology | 25 (14.0) | 2 (11.8) | 3 (20.0) | 8 (20.5) | 13 (12.4) | 9 (13.6) | 22 (22) | 8 (7.5) | 0 (0.0) | 28 (14.4) | 2 (12.5) | 30 (14.3) |
| Public health | 13 (7.3) | 0 (0.0) | 1 (6.7) | 3 (7.7) | 6 (5.7) | 5 (7.6) | 11 (11) | 3 (2.8) | 0 (0.0) | 12 (6.2) | 2 (12.5) | 14 (6.7) |
| Basic Science | 4 (2.2) | 0 (0.0) | 0 (0.0) | 1 (2.6) | 1 (1.0) | 2 (3.0) | 3 (3) | 1 (0.9) | 0 (0.0) | 4 (2.1) | 0 (0.0) | 4 (1.9) |
| *Have you been involved with monkeypox policy work?* | | | | | | | | | | | | |
| Yes | 66 (37.1) | 4 (23.5) | 7 (46.7) | 11 (28.2) | 38 (36.2) | 28 (42.4) | 42 (42) | 34 (32.1) | 1 (25.0) | 72 (37.1) | 5 (31.2) | 77 (36.7) |
| No | 112 (62.9) | 13 (76.5) | 8 (53.3) | 28 (71.8) | 67 (63.8) | 38 (57.6) | 58 (58) | 72 (67.9) | 3 (75.0) | 122 (62.9) | 11 (68.8) | 133 (63.3) |
| *Did you attend briefing meetings and calls with public health agencies?* | | | | | | | | | | | | |
| National | 45 (25.3) | 2 (11.8) | 3 (20.0) | 2 (5.1) | 29 (27.6) | 19 (28.8) | 25 (25.0) | 23 (21.7) | 2 (50.0) | 47 (24.2) | 3 (18.8) | 50 (23.8) |
| Regional | 63 (35.4) | 4 (23.5) | 1 (6.7) | 11 (28.2) | 34 (32.4) | 23 (34.8) | 35 (35.0) | 31 (29.2) | 2 (50.0) | 61 (31.4) | 7 (43.8) | 68 (32.3) |
| Local e.g. your clinic/service facility | 98 (55.1) | 12 (70.6) | 9 (60.0) | 28 (71.8) | 54 (51.4) | 37 (56.1) | 57 (57.0) | 62 (58.5) | 0 (0.0) | 110 (56.7) | 9 (56.2) | 119 (56.7) |
| International e.g. WHO, ECDC | 19 (10.7) | 0 (0.0) | 3 (20.0) | 3 (7.7) | 12 (11.4) | 7 (10.6) | 9 (9.0) | 12 (11.3) | 1 (25.0) | 19 (9.8) | 3 (18.8) | 22 (10.5) |
| None | 46 (25.8) | 3 (17.6) | 4 (26.7) | 6 (15.4) | 28 (26.7) | 19 (28.8) | 23 (23.0) | 30 (28.3) | 0 (0.0) | 51 (26.3) | 2 (12.5) | 53 (25.2) |
| Contributed to monkeypox research | Job title | | | Age in years | | | Gender | | | Ethnicity | | Total |
| | Consultant (n=61) | Doctor-in-training (n=9) | Nurse or AHP (n=7) | <35 (n=22) | 35-50 (n=29) | >50 (n=26) | Male (n=45) | Female (n=31) | Non-binary (n=1) | White (n=68) | All other groups (n=9) | (n=77) |
| *How much has your other research been affected as a result of your monkeypox research?* | | | | | | | | | | | | |
| Not at all | 16 (26.2) | 4 (44.4) | 3 (42.9) | 8 (36.4) | 4 (13.8) | 11 (42.3) | 15 (33.3) | 8 (25.8) | 0 (0.0) | 22 (32.4) | 1 (11.1) | 23 (29.9) |
| Suffered slightly | 17 (27.9) | 3 (33.3) | 3 (42.9) | 5 (22.7) | 12 (41.4) | 6 (23.1) | 12 (26.7) | 10 (32.3) | 1 (100.0) | 20 (29.4) | 3 (33.3) | 23 (29.9) |
| By a moderate amount | 13 (21.3) | 2 (22.2) | 0 (0.0) | 5 (22.7) | 5 (17.2) | 5 (19.2) | 10 (22.2) | 5 (16.1) | 0 (0.0) | 12 (17.6) | 3 (33.3) | 15 (19.5) |
| Considerably suffered | 13 (21.3) | 0 (0.0) | 1 (14.3) | 4 (18.2) | 6 (20.7) | 4 (15.4) | 7 (15.6) | 7 (22.6) | 0 (0.0) | 12 (17.6) | 2 (22.2) | 14 (18.2) |
| Extremely suffered | 2 (3.3) | 0 (0.0) | 0 (0.0) | 0 (0.0) | 2 (6.9) | 0 (0.0) | 1 (2.2) | 1 (3.2) | 0 (0.0) | 2 (2.9) | 0 (0.0) | 2 (2.6) |
| *Have you published or submitted any research to a journal on monkeypox during this outbreak?* | | | | | | | | | | | | |
| Yes | 35 (57.4) | 5 (55.6) | 3 (42.9) | 13 (59.1) | 16 (55.2) | 14 (53.8) | 26 (57.8) | 16 (51.6) | 1 (100.0) | 36 (52.9) | 7 (77.8) | 43 (55.8) |
| No | 26 (42.6) | 4 (44.4) | 4 (57.1) | 9 (40.9) | 13 (44.8) | 12 (46.2) | 19 (42.2) | 15 (48.4) | 0 (0.0) | 32 (47.1) | 2 (22.2) | 34 (44.2) |
| *Have you obtained grant money for research on monkeypox?* | | | | | | | | | | | | |
| Yes | 0 (0.0) | 1 (11.1) | 1 (14.3) | 1 (4.5) | 1 (3.4) | 0 (0.0) | 0 (0.0) | 1 (3.2) | 1 (100.0) | 0 (0.0) | 2 (22.2) | 2 (2.6) |
| No | 57 (93.4) | 8 (88.9) | 5 (71.4) | 20 (90.1) | 26 (89.7) | 24 (92.3) | 44 (97.8) | 26 (83.9) | 0 (0.0) | 63 (92.6) | 7 (77.8) | 70 (90.9) |
| Applied but not heard | 4 (6.6) | 0 (0.0) | 1 (14.3) | 1 (4.5) | 2 (6.9) | 2 (3.8) | 1 (8.9) | 4 (12.9) | 0 (0.0) | 5 (7.4) | 0 (0.0) | 5 (6.4) |
| *Have you been asked to be involved with any media outlets to do with monkeypox?* | | | | | | | | | | | | |
| Yes | 28 (45.9) | 2 (22.2) | 2 (28.6) | 5 (22.7) | 18 (62.1) | 9 (34.6) | 22 (48.9) | 9 (29.0) | 1 (100.0) | 28 (41.2) | 4 (44.4) | 32 (41.6) |
| No | 33 (54.1) | 7 (77.8) | 5 (71.4) | 17 (77.3) | 11 (37.9) | 17 (65.4) | 23 (51.1) | 22 (71.0) | 0 (0.0) | 40 (58.8) | 5 (55.6) | 45 (58.4) |
| Published or submitted any research | Job title | | | | | | Gender | | | Ethnicity | | Total |
| | Consultant (n=35) | Doctor-in-training (n=5) | Nurse or AHP (n=3) | <35 (n=13) | 35-50 (n=16) | >50 (n=14) | Men (n=26) | Women (n=16) | Non-binary (n=1) | White (n=36) | All other groups (n=7) | (n=43) |

| | | | | | | | | | | | | |
|---|---|---|---|---|---|---|---|---|---|---|---|---|
| *Did you collaborate with colleagues?* | | | | | | | | | | | | |
| In your own service | 11 (31.4) | 2 (40.0) | 2 (66.7) | 6 (46.2) | 6 (37.5) | 3 (21.4) | 9 (34.6) | 6 (37.5) | 0 (0.0) | 11 (30.6) | 4 (57.1) | 15 (34.8) |
| In your own country | 12 (34.3) | 0 (0.0) | 1 (33.3) | 3 (23.1) | 4 (25.0) | 6 (42.9) | 7 (26.9) | 6 (37.5) | 0 (0.0) | 12 (33.3) | 1 (14.3) | 13 (30.2) |
| In your own region | 4 (11.4) | 3 (60.0) | 0 (0.0) | 2 (15.4) | 4 (25.0) | 1 (7.1) | 4 (15.4) | 2 (12.5) | 1 (100.0) | 6 (16.7) | 1 (14.3) | 7 (16.3) |
| Globally | 8 (22.9) | 0 (0.0) | 0 (0.0) | 2 (15.4) | 2 (12.5) | 4 (28.6) | 6 (23.1) | 2 (12.5) | 0 (0.0) | 7 (19.4) | 1 (14.3) | 8 (18.6) |
| *What was your role within the research process?* | | | | | | | | | | | | |
| Collected data and named author | 15 (42.9) | 1 (20.0) | 0 (0.0) | 5 (38.5) | 4 (25.0) | 7 (50.0) | 12 (46.2) | 4 (25.0) | 0 (0.0) | 14 (38.9) | 2 (28.6) | 16 (37.2) |
| Collected data and part of a writing group | 13 (37.1) | 2 (40.0) | 3 (100.0) | 7 (53.8) | 7 (43.8) | 4 (28.6) | 7 (26.9) | 11 (68.8) | 0 (0.0) | 16 44.4) | 2 (28.6) | 18 (41.9) |
| Collected data only | 2 (5.7) | 1 (20.0) | 0 (0.0) | 0 (0.0) | 1 (6.3) | 2 (14.3) | 2 (7.7) | 0 (0.0) | 1 (100.0) | 1 (2.8) | 2 (28.6) | 3 (7.0) |
| Designed the study | 5 (14.3) | 1 (20.0) | 0 (0.0) | 1 (7.7) | 4 (25.0) | 1 (7.1) | 5 (19.2) | 1 (6.3) | 0 (0.0) | 5 (13.9) | 1 (14.3) | 6 (14.0) |

**Figure S1. Barchart comparing research active and not research active EU survery participants showing proportions by demographic characteristics, workplace, involvement in policy and public health agency (PHA) work.**

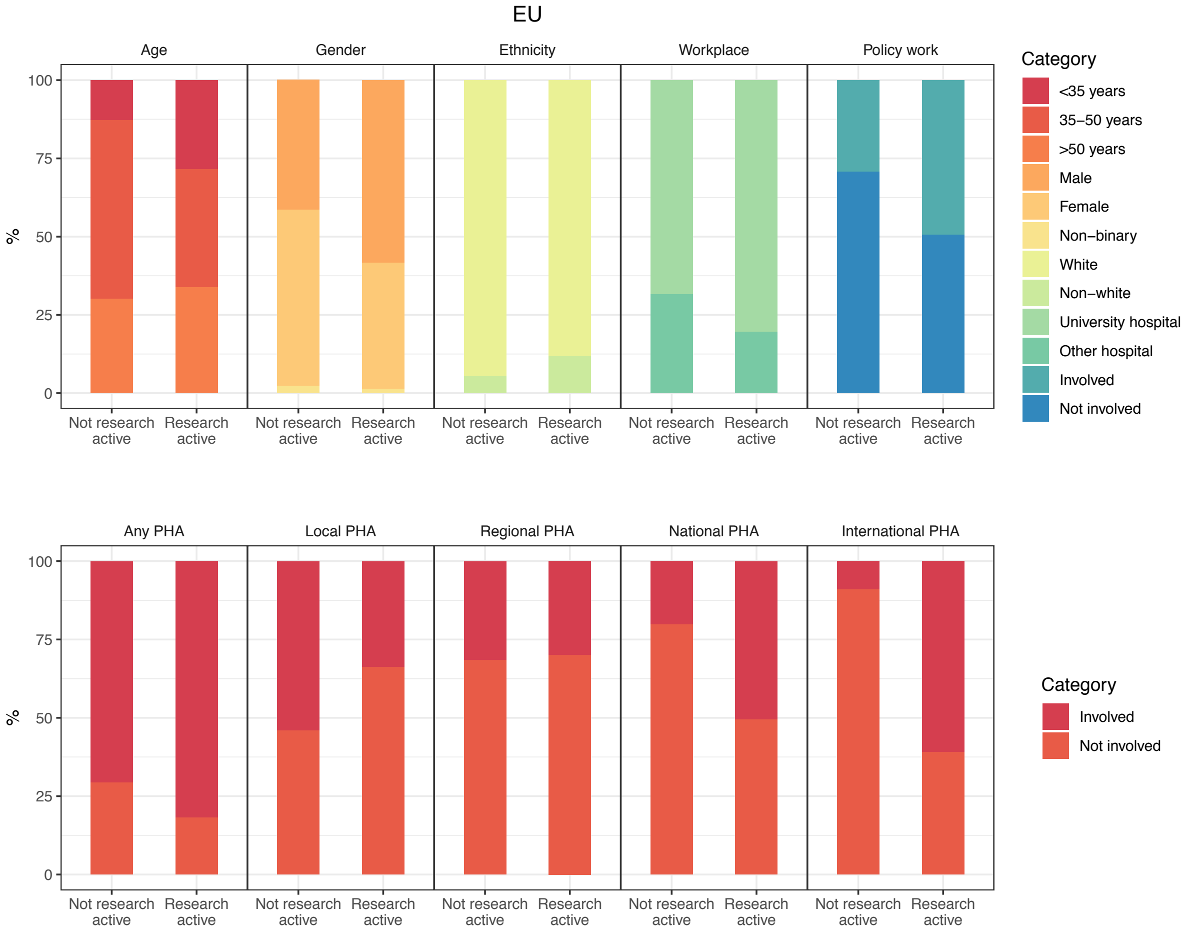

**Table S4. Summary of responses to research engagement questions from all US survey respondents. Results presented as n (%). AHP: allied health professional.**

| | Job title | | | Age in years | | | Gender | | | Ethnicity | | Total (n=58) |
|---|---|---|---|---|---|---|---|---|---|---|---|---|
| | Consultant (n=40) | Doctor-in-training (n=2) | Nurse or AHP (n=16) | <35 (n=8) | 35-50 (n=23) | >50 (n=27) | Male (n=27) | Female (n=30) | Non-binary (n=1) | White (n=38) | All other groups (n=20) | |
| *Have you contributed to monkeypox research?* | | | | | | | | | | | | |
| Yes | 16 (40.0) | 1 (50.0) | 5 (31.2) | 4 (50.0) | 11 (47.8) | 7 (25.9) | 9 (33.3) | 13 (43.3) | 0 (0.0) | 15 (39.5) | 7 (35.0) | 22 (37.9) |
| No | 24 (60.0) | 1 (50.0) | 11 (68.8) | 4 (50.0) | 12 (52.2) | 20 (74.1) | 18 (66.7) | 17 (56.7) | 1 (100.0) | 23 (60.5) | 13 (65.0) | 36 (62.1) |
| *How would you describe your area of research focus?* | | | | | | | | | | | | |
| Clinical | 16 (40.0) | 1 (50.0) | 5 (31.2) | 4 (50.0) | 11 (47.8) | 7 (25.9) | 9 (33.3) | 13 (43.3) | 0 (0.0) | 15 (39.5) | 7 (35.0) | 22 (37.5) |
| Epidemiology | 7 (17.5) | 0 (0.0) | 0 (0.0) | 1 (12.5) | 3 (13.0) | 3 (11.1) | 1 (3.7) | 6 (20.0) | 0 (0.0) | 4 (10.5) | 3 (15.0) | 7 (12.1) |
| Public health | 2 (5.0) | 0 (0.0) | 1 (6.2) | 0 (0.0) | 2 (8.7) | 1 (3.7) | 0 (0.0) | 3 (10.0) | 0 (0.0) | 3 (7.9) | 0 (0.0) | 3 (5.2) |
| Basic Science | 1 (2.5) | 0 (0.0) | 0 (0.0) | 0 (0.0) | 1 (4.3) | 0 (0.0) | 0 (0.0) | 1 (3.3) | 0 (0.0) | 0 (0.0) | 1 (5.0) | 1 (1.7) |
| *Have you been involved with monkeypox policy work?* | | | | | | | | | | | | |
| Yes | 14 (35.0) | 0 (0.0) | 4 (25.0) | 2 (25.0) | 7 (30.4) | 9 (33.3) | 9 (33.3) | 9 (30.0) | 0 (0.0) | 12 (31.6) | 6 (30.0) | 18 (31.0) |
| No | 26 (65.0) | 2 (100.0) | 12 (75.0) | 6 (75.0) | 26 (69.6) | 18 (66.7) | 18 (66.7) | 21 (70.0) | 1 (100.0) | 26 (68.4)) | 14 (70.0) | 40 (69.0) |
| *Did you attend briefing meetings and calls with public health agencies?* | | | | | | | | | | | | |
| National | 12 (30.0) | 0 (0.0) | 2 (12.5) | 2 (25.0) | 5 (21.7) | 7 (25.9) | 7 (25.9) | 7 (23.3) | 0 (0.0) | 6 (15.8) | 8 (40.0) | 14 (24.1) |
| Regional | 14 (35.0) | 0 (0.0) | 6 (37.5) | 1 (12.5) | 7 (30.4) | 12 (44.4) | 6 (22.2) | 14 (46.7) | 0 (0.0) | 15 (39.5) | 5 (25.0) | 20 (34.5) |
| Local e.g. your clinic/service facility | 25 (62.5) | 2 (100.0) | 11 (68.8) | 7 (87.5) | 19 (82.6) | 12 (44.4) | 16 (59.3) | 22 (73.3) | 0 (0.0) | 24 (63.2) | 14 (70.0) | 38 (65.5) |
| International e.g. WHO, ECDC | 5 (12.5) | 0 (0.0) | 0 (0.0) | 1 (12.5) | 1 (4.3) | 3 (11.1) | 3 (11.1) | 2 (6.7) | 0 (0.0) | 2 (5.3) | 3 (15.0) | 5 (8.6) |
| None | 9 (22.5) | 0 (0.0) | 5 (31.2) | 1 (12.5) | 4 (17.4) | 9 (33.3) | 8 (29.6) | 5 (16.7) | 1 (100.0) | 10 (26.3) | 4 (20.0) | 14 (24.1) |
| Contributed to monkeypox research | Job title | | | Age in years | | | Gender | | | Ethnicity | | Total (n=22) |
| | Consultant (n=16) | Doctor-in-training (n=1) | Nurse or AHP (n=5) | <35 (n=4) | 35-50 (n=11) | >50 (n=7) | Male (n=9) | Female (n=13) | Non-binary (n=0) | White (n=15) | All other groups (n=7) | |
| *How much has your other research been affected as a result of your monkeypox research?* | | | | | | | | | | | | |
| Not at all | 7 (43.8) | 0 (0.0) | 1 (20.0) | 2 (50.0) | 3 (27.3) | 3 (42.9) | 3 (33.3) | 5 (38.5) | - | 6 (40.0) | 2 (28.6) | 8 (36.3) |
| Suffered slightly | 7 (43.8) | 1 (100.0) | 2 (40.0) | 2 (50.0) | 4 (36.4) | 4 (57.1) | 4 (44.4) | 6 (46.2) | - | 7 (46.7) | 3 (42.9) | 10 (45.5) |
| By a moderate amount | 1 (6.3) | 0 (0.0) | 2 (40.0) | 0 (0.0) | 3 (27.3) | 0 (0.0) | 2 (22.2) | 1 (7.7) | - | 1 (6.7) | 2 (28.6) | 3 (13.6) |
| Considerably suffered | 0 (0.0) | 0 (0.0) | 0 (0.0) | 0 (0.0) | 0 (0.0) | 0 (0.0) | 0 (0.0) | 0 (0.0) | - | 0 (0.0) | 0 (0.0) | 0 (0.0) |
| Extremely suffered | 1 (6.3) | 0 (0.0) | 0 (0.0) | 0 (0.0) | 1 (9.1) | 0 (0.0) | 0 (0.0) | 1 (7.7) | - | 1 (6.7) | 0 (0.0) | 1 (4.5) |
| *Have you published or submitted any research to a journal on monkeypox during this outbreak?* | | | | | | | | | | | | |
| Yes | 5 (31.3) | 1 (100.0) | 0 (0.0) | 2 (50.0) | 2 (18.2) | 2 (28.6) | 1 (11.1) | 5 (38.5) | - | 4 (26.7) | 2 (28.6) | 6 (27.3) |
| No | 11 (68.8) | 0 (0.0) | 5 (100.0) | 2 (50.0) | 9 (81.8) | 5 (71.4) | 8 (88.9) | 8 (61.5) | - | 11 (73.3) | 5 (71.4) | 16 (72.7) |
| *Have you obtained grant money for research on monkeypox?* | | | | | | | | | | | | |
| Yes | 4 (25.0) | 0 (0.0) | 0 (0.0) | 0 (0.0) | 3 (27.3) | 1 (14.3) | 2 (22.2) | 2 (15.4) | - | 2 (13.3) | 2 (28.6) | 4 (18.1) |
| No | 10 (62.5) | 1 (100.0) | 5 (100.0) | 4 (100.0) | 8 (72.8) | 4 (57.1) | 6 (66.7) | 10 (76.9) | - | 12 (80.0) | 4 (57.1) | 16 (72.7) |
| Applied but not heard | 2 (12.5) | 0 (0.0) | 0 (0.0) | 0 (0.0) | 0 (0.0) | 2 (28.6) | 1 (11.1) | 1 (7.7) | - | 1 (6.7) | 1 (14.3) | 2 (9.1) |
| *Have you been asked to be involved with any media outlets to do with monkeypox?* | | | | | | | | | | | | |
| Yes | 10 (62.5) | 0 (0.0) | 0 (0.0) | 0 (0.0) | 5 (45.5) | 5 (71.4) | 4 (44.4) | 6 (46.2) | - | 7 (46.7) | 3 (42.9) | 10 (45.5) |
| No | 6 (37.5) | 1 (100.0) | 5 (100.0) | 4 (100.0) | 6 (54.5) | 2 (28.6) | 5 (55.6) | 7 (53.8) | - | 8 (53.3) | 4 (57.1) | 12 (54.5) |
| Published or submitted any research | Job title | | | | | | Gender | | | Ethnicity | | Total (n=6) |
| | Consultant (n=5) | Doctor-in-training (n=1) | Nurse or AHP (n=0) | <35 (n=2) | 35-50 (n=2) | >50 (n=2) | Men (n=1) | Women (n=5) | Non-binary (n=0) | White (n=4) | All other groups (n=2) | |

| | | | | | | | | | | | | |
|---|---|---|---|---|---|---|---|---|---|---|---|---|
| *Did you collaborate with colleagues?* | | | | | | | | | | | | |
| In your own service | 0 (0.0) | 0 (0.0) | - | 0 (0.0) | 0 (0.0) | 0 (0.0) | 0 (0.0) | 0 (0.0) | - | 0 (0.0) | 0 (0.0) | 0 (0.0) |
| In your own country | 3 (60.0) | 1 (100.0) | - | 2 (100.0) | 1 (50.0) | 1 (50.0) | 1 (100.0) | 3 (60.0) | - | 3 (75.0) | 1 (50.0) | 4 (66.7) |
| In your own region | 1 (20.0) | 0 (0.0) | - | 0 (0.0) | 0 (0.0) | 1 (50.0) | 0 (0.0) | 1 (20.0) | - | 1 (25.0) | 0 (0.0) | 1 (16.7) |
| Globally | 1 (20.0) | 0 (0.0) | - | 0 (0.0) | 1 (50.0) | 0 (0.0) | 0 (0.0) | 1 (20.0) | - | 0 (0.0) | 1 (50.0) | 1 (16.7) |
| *What was your role within the research process?* | | | | | | | | | | | | |
| Collected data and named author | 3 (60.0) | 0 (0.0) | - | 0 (0.0) | 2 (100.0) | 1 (50.0) | 0 (0.0) | 3 (60.0) | - | 1 (25.0) | 2 (100.0) | 3 (50.0) |
| Collected data and part of a writing group | 1 (20.0) | 1 (100.0) | - | 1 (50.0) | 0 (0.0) | 1 (50.0) | 0 (0.0) | 2 (40.0) | - | 2 (50.0) | 0 (0.0) | 2 (33.3) |
| Collected data only | 0 (0.0) | 0 (0.0) | - | 0 (0.0) | 0 (0.0) | 0 (0.0) | 0 (0.0) | 0 (0.0) | - | 0 (0.0) | 0 (0.0) | 0 (0.0) |
| Designed the study | 1 (20.0) | 0 (0.0) | - | 1 (50.0) | 0 (0.0) | 0 (0.0) | 1 (100.0) | 0 (0.0) | - | 1 (25.0) | 0 (0.0) | 1 (16.7) |

**Figure S2. Barchart comparing research active and not research active US survery participants showing proportions by demographic characteristics, workplace, involvement in policy and public health agency (PHA) work.**

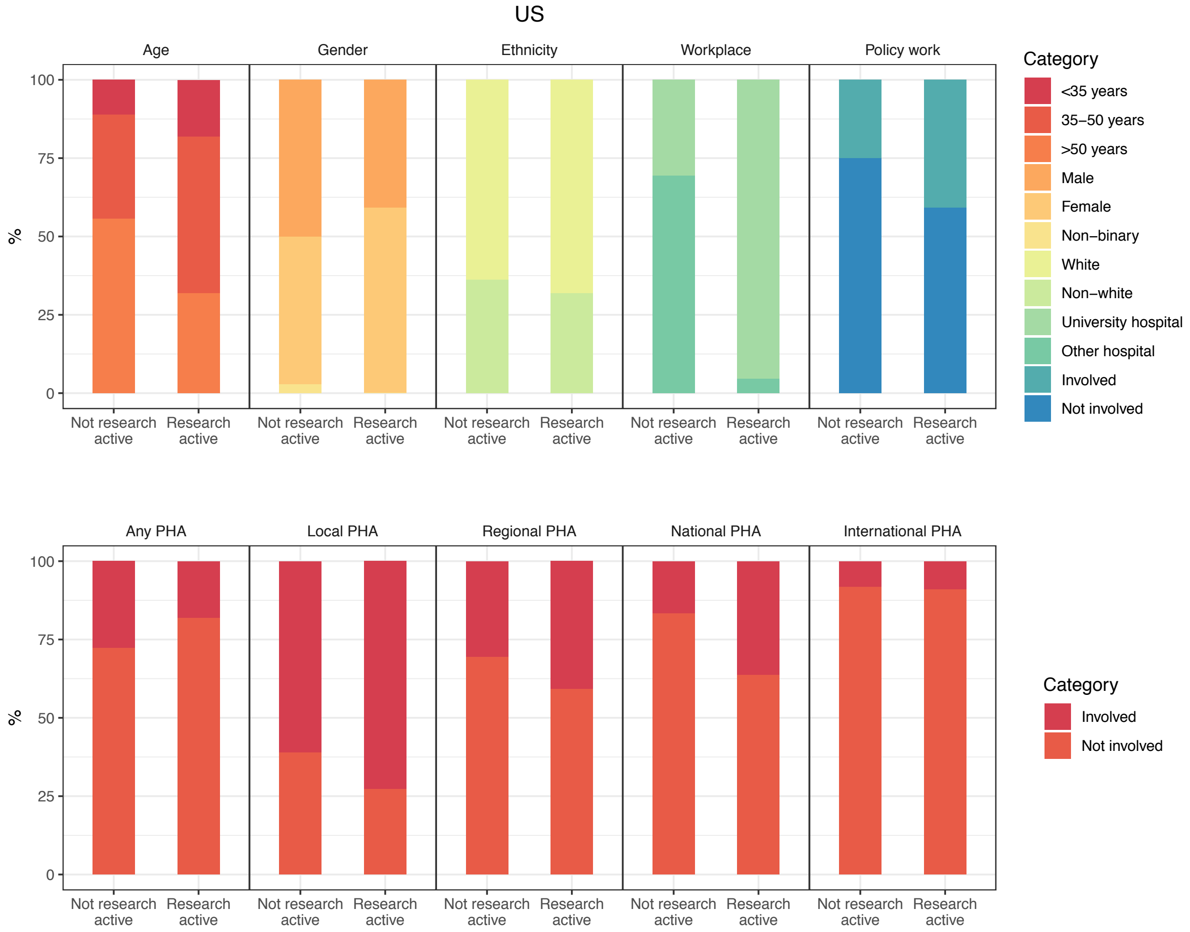
